# Objective measures of reward sensitivity and motivation in people with high vs low anhedonia

**DOI:** 10.1101/2021.10.21.21265287

**Authors:** Chloe Slaney, Adam M. Perkins, Robert Davis, Ian Penton-Voak, Marcus R Munafò, Conor J. Houghton, Emma S.J. Robinson

## Abstract

**Background:** Anhedonia – a diminished interest or pleasure in activities – is a core self-reported symptom of depression which is poorly understood and often resistant to conventional antidepressants. This symptom may occur due to dysfunction in one or more sub-components of reward processing: motivation, consummatory experience, and/or learning. However, the precise impairments remain elusive. Dissociating these components (ideally, using cross-species measures) and relating them to the subjective experience of anhedonia is critical as it may benefit fundamental biology research and novel drug development.

**Methods:** Using a battery of behavioural tasks based on rodent assays, we examined reward motivation (Joystick-Operated Runway Task, JORT; and Effort-Expenditure for Rewards Task, EEfRT) and reward sensitivity (Sweet Taste Test) in a non-clinical population who scored high (*N* = 32) or low (*N* = 34) on an anhedonia questionnaire (Snaith-Hamilton Pleasure Scale).

**Results:** Compared to the low anhedonia group, the high anhedonia group displayed marginal impairments in effort-based decision-making (EEfRT) and reduced reward sensitivity (Sweet Taste Test). However, we found no evidence of a difference between groups in physical effort exerted for reward (JORT). Interestingly, whilst the EEfRT and Sweet Taste Test correlated with anhedonia measures, they did not correlate with each other. This poses the question of whether there are subgroups within anhedonia; however, further work is required to directly test this hypothesis.

**Conclusions:** Our findings suggest that anhedonia is a heterogenous symptom associated with impairments in reward sensitivity and effort-based decision-making.

---

Anhedonia, a markedly diminished interest or pleasure in activities (DSM-5; American Psychiatric Association, 2013), is a core self-reported symptom of Major Depressive Disorder (MDD) that responds poorly to conventional treatments (Uher et al., 2012). Despite its importance, the behavioural and neurobiological basis of anhedonia remains poorly understood (Cooper, Arulpragasam, & Treadway, 2018). This could, in part, be due the over-reliance on questionnaires to measure this symptom. Objective measures, particularly those that can be applied in both clinical and preclinical animal research, may help to improve our mechanistic understanding of anhedonia and aid the development of targeted treatments (Der-Avakian & Markou, 2012; Pizzagalli, Jahn, & O’Shea, 2005; Thomsen, 2015; Treadway & Zald, 2011).

One way to objectively measure anhedonia is behaviourally. Anhedonia may occur due to dysfunction in one or more components of reward processing: motivation to obtain rewards (“wanting”), consumption of rewards (“detection” or “liking”), and learning what predicts rewards (“learning”) (Berridge & Robinson, 2003). Whilst the taxonomy of reward processing is debated, preclinical studies suggest that these sub-domains involve partially dissociable neural systems (Berridge & Robinson, 1998; Berridge et al., 2009). Despite advancements in preclinical research (Der-Avakian et al., 2013), few studies have tried to assess these different components in people and relate them to questionnaire measures of anhedonia (McCabe, 2018). Translating behavioural tasks from animals to humans and assessing which components relate to anhedonia is critical as it may provide an objective biomarker that could be used in the development and testing of new treatments (Der-Avakian, Barnes, Markou, & Pizzagalli, 2016). As most preclinical tasks focus on measuring reward motivation and consumption, we also focus on these two components here.

Reward motivation is the “incentive or desire to act or accomplish goals” (Der-Avakian et al., 2016). Recently, preclinical reward motivation tasks, such as effort-related choice tasks (Salamone, Correa, Yang, Rotolo, & Presby, 2018) and the Progressive Ratio Task (Hodos, 1961), have been adapted for human studies (Hershenberg et al., 2016; Treadway, Buckholtz, Schwartzman, Lambert, & Zald, 2009). For example, the Effort-Expenditure for Rewards Task (EEfRT; Treadway et al., 2009), requires participants to choose whether to exert more effort for a high reward or less effort for a low reward over many trials. Interestingly, people with MDD and people who have higher levels of anhedonia in a non-clinical population choose the high effort/high reward option less often than healthy controls suggesting impairments in effort-based decision-making (Treadway, Bossaller, Shelton, & Zald, 2012; Treadway et al., 2009). In the Progressive Ratio Task, incrementally more effortful responses are required to obtain the same amount of reward, and motivation is operationalised as the point at which the individual gives up (i.e., their “breakpoint”) (Strauss et al., 2016). In this task, people with MDD give up earlier than healthy controls (Hershenberg et al., 2016), suggesting a reduced willingness to work for reward. Whilst there is accumulating evidence showing reduced reward motivation in MDD, few studies focus on physical effort-expenditure for reward (Halahakoon et al., 2020) (although see Cléry-Melin et al., 2011). To address this gap, we developed a novel reward motivation task (Joystick-Operated Runway Task; JORT; Perkins et al., 2009), designed to measure a person’s physical effort exerted for reward.

Reward sensitivity is the consummatory experience of reward. Here, we use the term reward sensitivity to refer to reward detection, not reward liking which is how it is often used in the human literature. We focus on reward detection, rather than reward liking, to increase the back translation of the results to the animal work. The most commonly used reward sensitivity task in rodents is the sucrose preference test (SPT; Willner et al., 1987), which measures an animal’s ability to detect, and show a preference for, a weak sucrose solution over water. Researchers have attempted to translate this task into humans using a Sweet Taste Test (Amsterdam, Settle, Doty, Abelman, & Winokur, 1987; Berlin, Givry-Steiner, Lecrubier, & Puech, 1998; Dichter, Smoski, Kampov-Polevoy, Gallop, & Garbutt, 2010). Although different variations have been used, they typically require participants to report the intensity and pleasantness of different sucrose concentrations. Previous studies have reported similar pleasantness ratings of sucrose in people with MDD compared to healthy controls (Amsterdam et al., 1987; Berlin et al., 1998; Dichter et al., 2010). However, there is discrepancy across studies when measuring sucrose intensity: some studies have reported poorer detection of sucrose in people with MDD compared to healthy controls (Amsterdam et al., 1987; Berlin et al., 1998), whilst others have failed to find any differences (Dichter et al., 2010). One potential explanation is that point scales, often used to measure ability to detect sucrose, may not be sensitive enough to reliably detect impairments (McCabe, 2018).

Despite progress in the development of translational measures (Thomsen, 2015), there are gaps in the literature. First, most studies examine only one sub-component of reward (e.g., motivation) using one assay. However, a battery of tasks designed to probe different reward components (e.g., motivation, sensitivity) can demonstrate whether different components can be dissociated and which components are associated with anhedonia (Husain & Roiser, 2018; Nielson et al., 2020). Second, most studies compare people with MDD to healthy controls, with few directly measuring anhedonia using anhedonia questionnaires (McCabe, 2018). This is surprising, as only ∼ 37% people with MDD exhibit significant levels of anhedonia (Pelizza & Ferrari, 2009). In line with the Research Domain Criteria (RDoC), a symptom-based approach which examines behavioural tasks in relation to symptom-based questionnaires may provide more rigorous findings (Cuthbert, 2014; Insel et al., 2010).

Here, we compared individuals with high versus low levels of anhedonia in a non-clinical population (Snaith et al., 1995) on a battery of behavioural tasks designed to measure reward motivation (EEfRT and JORT) and reward sensitivity (Sweet Taste Test). A novel measure of reward motivation - the JORT - was designed to assess reward motivation in the absence of explicit decision-making. These tasks were chosen based on their potential to dissociate reward components and their potential for cross-species research. Based on previous research, we predicted that compared to people with low levels of anhedonia, people with high levels of anhedonia will show reduced reward motivation and reward sensitivity.

## Method

This study was pre-registered on the Open Science Framework (https://osf.io/p4zt6) and was approved by the Faculty of Biomedical Sciences Research Ethics Committee at the University of Bristol (Ref: 74082).

### Screening

Individuals who scored high (score ≥ 25) or low (score < 18) on the Snaith-Hamilton Pleasure Scale (SHAPS; see Figure 1)(Snaith et al., 1995) in a non-clinical population were invited to take part in this study. These individuals scored in the top and bottom 25^th^ percentile on the SHAPS, which was administered online to 380 volunteers. In the pre-registration, the original cut-off was a low score of < 17. However, due to initial difficulties with recruitment, this was increased to < 18 during the study.

**Figure 1.**
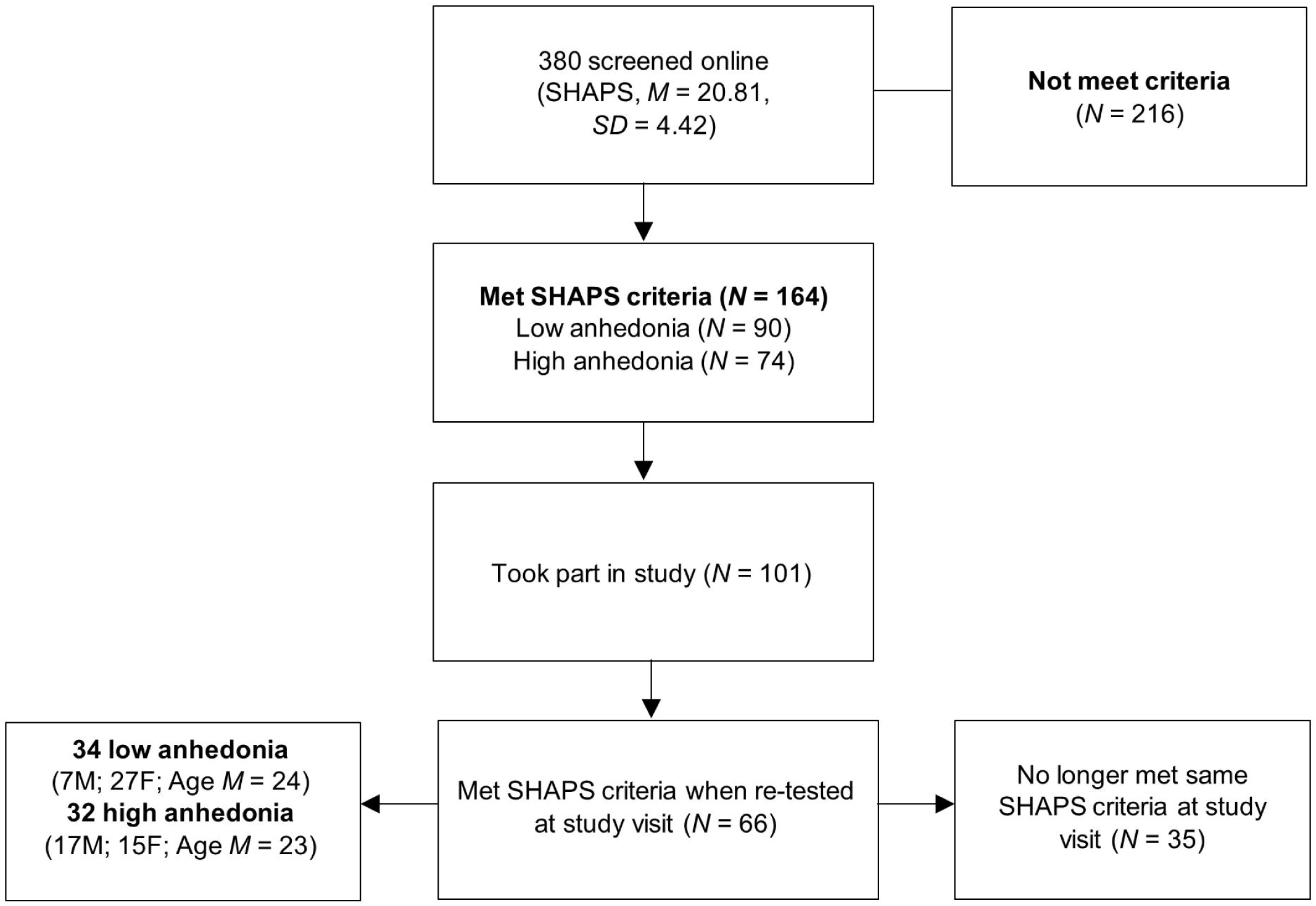
CONSORT diagram of study selection process.

### Participants

Whilst 101 participants (44 low anhedonia, 57 high anhedonia) met the online SHAPS criteria and took part in the study, only 66 (34 low, 32 high) still met their SHAPS criteria when re-tested at the study visit and were included in the primary analysis. For sample size calculations, see supplementary materials.

Eligibility criteria were aged ≥ 18 years, fluent in English, not suffering from a mental health condition or neurological illness (self-report), no current physical injuries, no allergy or intolerance to sugar, no disorder of taste or smell, not diabetic, not previously participated in a study using the JORT.

Participants were recruited using advertisements within the University of Bristol and volunteer databases. Participants were informed that they would receive £20 for their time (∼ 2 hours) and an additional performance-based pay (up to £5) based on one of the two reward motivation tasks (chosen randomly).

### Behavioural Measures

#### Joystick-Operated Runway Task

The JORT was originally developed as a translational measure of fear and anxiety, here it has been adapted as a measure of reward motivation. This is a computerised task which uses a force-sensing joystick to measure a person’s physical effort to obtain a reward (Perkins et al., 2009; PH-JS1, Psyal, London). Each trial begins with a green dot (representing the participant) positioned along an onscreen runway. A cue in the top left-hand corner of the screen indicates the number of points on offer (0, 10, 100 or 1,000 points; see Figure 2). Following this, a target (black dot) appears and immediately accelerates away from the participant. Participants are informed that to win the points on offer, they must push the joystick to chase and catch the onscreen target. The speed of the participants cursor is linked to the force applied on the joystick (Perkins et al., 2009). Trials vary in the number of points on offer (0, 10, 100 and 1,000 points; visible to the participant) and the minimum effort required to win (50, 80, 100 or 120% of their maximum calibrated force; not visible). If the participant catches the target, they receive feedback that they have won the number of points on offer. Trials end either upon catching the target or after 7 seconds. The inter-trial interval varies pseudo-randomly between 3-7 seconds. After each trial, the next trial automatically begins. Force applied to the joystick is recorded every 15 ms.

**Figure 2.**
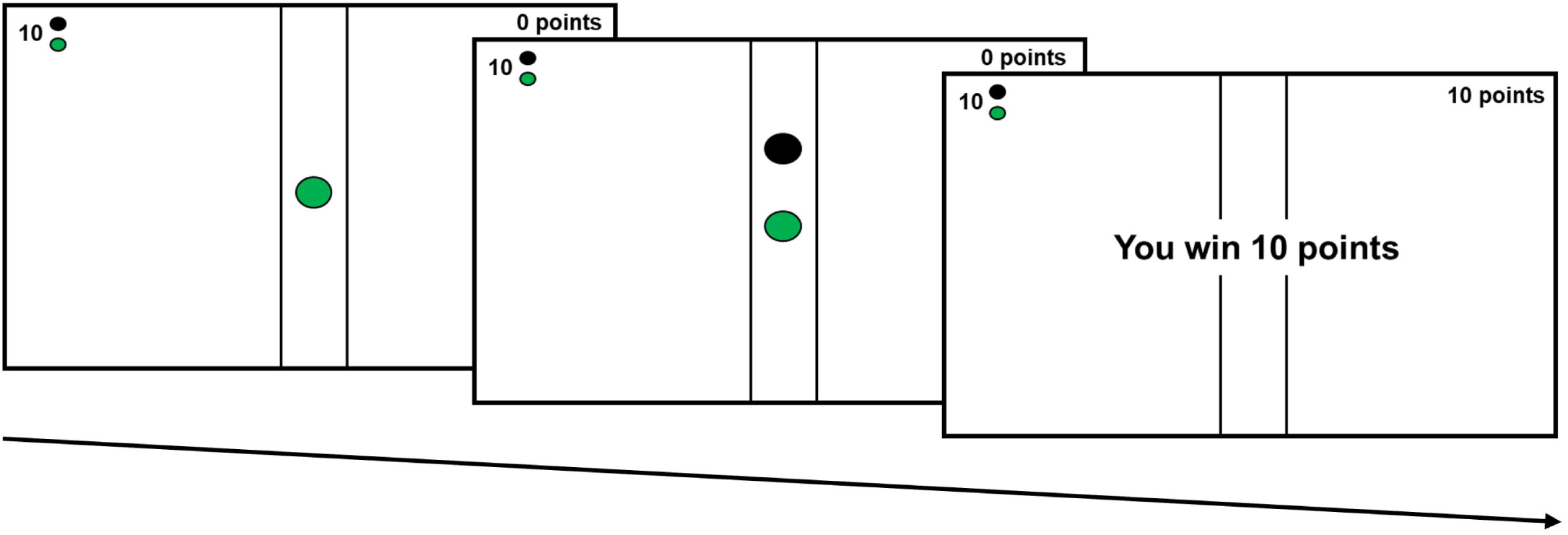
Example trial on the JORT. The green dot (representing the participant) is initially displayed along with a cue indicating the number of points on offer. This is followed by the presentation of the target (a black dot). The participant then receives feedback if they catch the target (e.g., "You win 10 points") or no feedback if they fail to catch the target.

Participants complete two blocks of 48 trials (96-total), separated by a short break (∼ 5 minutes). Each block contains an equal number of reward-effort combinations (*N* = 3), except for one combination where two trials were presented due to experimenter error (0 points-80% effort in second block). All participants complete the same pre-randomised order of trials. For a proof-of-concept study of the JORT, see supplementary materials.

Prior to the task, participants are given standardised verbal instructions before completing a calibration phase and four practice trials. In the calibration phase, participants push the joystick as “hard as they can” five times whenever “GO” is presented on the screen. Peak force reached during the calibration or practice trials (whichever is highest) is recorded as a person’s max calibrated force. To standardise the amount of effort required across participants on the main task, effort levels reflect the percentage of a participants max calibrated force.

#### Effort-Expenditure for Rewards Task

The EEfRT (Treadway et al., 2009) is a computerised effort-based decision-making task. On each trial, participants choose between an “easy” and “hard” option. The easy option requires 30 button presses within 7 seconds (using the index finger on their dominant hand) for 50p. The hard option requires 98 button presses (using the little finger on their non-dominant hand) within 21 seconds for 58p - £2. Participants have 5 seconds to choose, or they are randomly assigned to either option for that trial. Each trial also contains a visible probability cue (12%, 50%, 88%) which indicates the probability of winning money if they successfully complete the trial (i.e., win trials). The probability cue applies to both the easy and hard option, with an equal proportion across the experiment. Participants are informed that two “win trials” may be paid to them at the end of the experiment (performance-based pay). Participants complete the same pre-randomised order of trials for 20 minutes. For further details, see supplementary materials.

#### Sweet Taste Test

We employed a modified version of the Sweet Taste Test (Berlin et al., 1998), which measures detection threshold of sweet taste. On each trial, participants were given one sucrose concentration. They were asked to sip one mouthful of the solution, swish it around their mouth for 5 seconds and then spit it out (Berlin et al., 1998). Participants were asked if they could detect the presence of sugar in the solution (Berlin et al., 1998). In between trials, participants sipped one mouthful of water, swished it around their mouth for 5 seconds and then spat this out. Prior to the task, participants completed two practice trials containing water to ensure that they were familiar with the procedure and to clean their palette. Seven concentrations of sucrose were used (0, 0.5, 1, 1.5, 2, 2.5, 5% w/v), each delivered in 15 mL. A staircase method with five reversals was employed. The first concentration administered was always the strongest concentration (5% sucrose) to ensure participants were aware of what to detect. A staircase method was used as it provides a fast determination of threshold. The detection threshold was calculated as the mean of the five boundaries (i.e., points at which they changed response from either detect to fail to detect, or vice versa).

### Self-Report Measures

The primary measure of anhedonia was the SHAPS (Snaith et al., 1995). This is a 14-item questionnaire which asks participants to report their ability to experience pleasure in the last few days. Additional measures of anhedonia, apathy and depression were collected: Chapman Physical Anhedonia Scale (CPAS; Chapman et al., 1976), Temporal Experience of Pleasure Scale (TEPS; Gard et al., 2006), Beck Depression Inventory-II (BDI-II; Beck et al., 1996) and Apathy Evaluation Scale (AES; Marin et al., 1991). Higher scores indicate higher symptoms on all questionnaires except for the TEPS. For further details, see supplementary materials.

### Procedure

Participants completed the behavioural tasks in the following order: EEfRT, JORT and the Sweet Taste Test, which took approximately one hour. Participants then completed the questionnaires: demographic form, SHAPS, CPAS, TEPS, BDI-II, AES.

### Data Analysis

#### Primary Analysis

Data were analysed using SPSS 24 (IBM). Primary analyses were run on those who met the SHAPS cut-offs at the study visit (*N* = 66). Based on previous research, *a priori* potential co-variates were examined and their main effects retained in the model (Engqvist, 2005).

For the JORT, 2 × 4 (group x reward) mixed ANOVAs examined differences between groups in relative average force, maximum force, and reaction time. The primary outcome was relative average force. For the EEfRT, 2 × 3 (group x probability) mixed ANCOVAs examined differences between groups in mean proportion of hard-task choices and reaction time. Sex was included as a covariate (Treadway et al., 2009). The primary outcome was proportion of hard-task choices. For the Sweet Taste Test, an ANCOVA examined differences between groups in detection threshold. Current smoking status (3 levels: daily, less than daily, not at all) was included as a covariate (Sato, Endo, & Tomita, 2002). All secondary outcomes and data checks are reported in the supplementary materials. Due to the high and low anhedonia group not being well-matched regarding sex, sensitivity analyses were run including sex as a covariate to ensure that any findings were not being driven by sex differences.

#### Exploratory Analysis

Spearman correlations examined associations between tasks and self-report measures, and correlations between different tasks. As these correlational analyses were exploratory, we report them descriptively only.

## Results

Data are available at the University of Bristol data repository, data.bris, at https://doi.org/10.5523/bris.1wlrhv4jzqs7q2egf8i7ruzta1.

### Participant Characteristics

For participant demographics, see Table 1. For distribution of scores on self-report measures and correlations between measures, see Supplementary Materials.

**Table 1.** Demographic data and questionnaire scores for participants in the high and low anhedonia group.

|  | Low Group |  | High Group |  | Group Difference |  |  |
| --- | --- | --- | --- | --- | --- | --- | --- |
|  | Mean | SD / % | Mean | SD / % | *F* | *p*-value | Post-hoc |
| Age | 24.0 | 5.1 | 22.8 | 8.6 | .41 | .52 | n/a |
| Males (*N*) | 7 | 21% | 17 | 53% | $X^2 = 7.54$ | .006 | H > L |
| Smoke Status | 2.7 | 0.7 | 2.7 | 0.6 | $X^2 = .23$ | .89 | n/a |
| Income | 2.3 | 0.6 | 2.2 | 0.6 | $X^2 = .64$ | .73 | n/a |
| Money Concern | 12 | 36% | 16 | 50% | $X^2 = 1.23$ | .27 | n/a |
| SHAPS | 15.4 | 1.0 | 27.6 | 2.2 | 838.34 | <.001 | H > L |
| CPAS | 9.6 | 5.7 | 18.0 | 7.1 | 27.67 | <.001 | H > L |
| TEPS – A | 50.5 | 4.6 | 40.3 | 5.2 | 71.10 | <.001 | L > H |
| TEPS – C | 40.6 | 5.3 | 30.3 | 6.2 | 52.80 | <.001 | L > H |
| BDI-II | 6.4 | 5.5 | 11.4 | 7.6 | 9.41 | .003 | H > L |
| Apathy Scale | 24.6 | 4.4 | 32.4 | 6.2 | 33.74 | <.001 | H > L |

*Note.* Current smoke status (1=daily, 2=less than daily, 3=not at all); income (1=high, 2=medium, 3=low); money concern, participants who reported “yes” to finance concerns; SHAPS, Snaith-Hamilton Pleasure Scale; TEPS, Temporal Experience of Pleasure Scale (A, anticipatory; C, consummatory); BDI-II, Beck Depression Inventory-II; n/a, not applicable. Welch’s ANOVA and Chi Square to compare groups.

### Joystick-Operated Runway Task

Based on *a priori* criteria, participants who succeeded in over 75% trials (*N* = 4) were excluded from the analysis. This is because these participants must have achieved at least one trial designed to be impossible (120% effort trials) and were therefore considered to have not successfully achieved their maximum force during the calibration. In total, 62 participants (33 low anhedonia, 29 high anhedonia) were included in the analysis.

#### Average force

There was strong evidence of a main effect of reward, participants exerted more force for higher reward magnitudes, *F*_(1.45,87.14)_ = 39.41, *p* <.001, n_p_^2^ = .40, Figure 3. Bonferroni-corrected pairwise comparisons revealed evidence of a difference between all reward magnitudes (*p*s ≤ .003). There was no evidence of a main effect of group (*F*_(1,60)_ = .004, *p* = .95, n_p_^2^ < .001; see Figure 3) or reward x group interaction (*F*_(1.45,87.14)_ = .56, *p* = .52, n_p_^2^= .009). In the sensitivity analyses, where sex was added to the model, there was no evidence of a main effect of sex (*p* = .58), group (*p* = .80) or reward x group interaction (*p* = .66).

**Figure 3.**
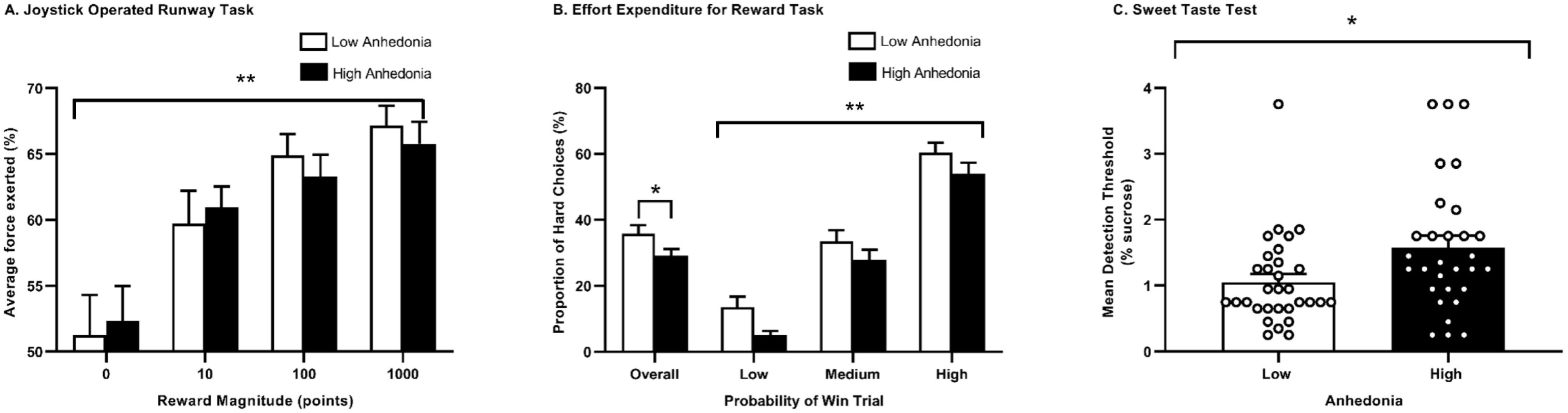
Differences between the high and low anhedonia group in the (A) Joystick-Operated Runway Task: relative average force exerted for each reward magnitude (*N* = 33 low, 29 high) (B) Effort-Expenditure for Rewards Task: mean proportion of hard-task choices across different levels of probability (*N* = 34 low, 32 high) and (C) Sweet Taste Test (*N* = 31 low, 30 high). Points represent participants. Error bars represent SEM. Data presented with statistical outliers included. * *p* < .05, ** *p* < .001.

### Effort-Expenditure for Rewards Task

Consistent with Treadway (2009), an upper bound of the first 50 trials was used in the analysis. Trials where participants did not make a choice were not included in the analysis (*M* = 49.20, *SD* = 1.90, range = 39 to 50).

#### Proportion of hard-task choices

There was strong evidence of a main effect of probability, participants chose the hard-task choice more often when the probability of winning was higher, *F*_(2,126)_ = 209.12, *p* < .001, n_p_^2^ = .77, Figure 3. Bonferroni-corrected comparisons revealed evidence of a difference between all probabilities (*p*s < .001). There was evidence of a main effect of group, *F*_(1,63)_ = 4.45, *p* = .039, n_p_^2^ = .07: the high anhedonia group (*M* = 28.88, *SE* = 2.42) chose the hard-task choice less often than the low anhedonia group (*M* = 36.41, *SE* = 2.58; see Figure 3). There was no evidence of other main effects or interactions (*p*s ≥ .57). However, removal of one outlier weakened the evidence of the main effect of group, *F*_(1,62)_ = 2.64, *p* =.109, n_p_^2^ = .041). Independent *t*-tests indicated modest evidence of a difference between groups on low probability trials (*p* = .016), with no clear evidence of a difference on medium or high probability trials (*p*s ≥ .16).

#### Button Pressing Rate

There was no evidence of a difference between groups in mean button pressing rate (button-pressing speed) for the easy or hard-task choices (*p*s ≥ .11).

### Sweet Taste Test

Based on *a priori* criteria, participants who reported detection of sucrose in 0% sucrose concentrations were excluded. This excluded 5 participants; 61 participants (31 low anhedonia, 30 high anhedonia) were included in the analysis.

The analysis was conducted on log transformed data, which met ANOVA assumptions. There was evidence of a main effect of group, the high anhedonia group required a higher concentration of sucrose to detect its presence, *F*_(1,57)_ = 4.56, *p* = .037, n_p_^2^ = .07, see Figure 3. There was no evidence of a main effect of smoking status, *F*_(2,57)_ = .41, *p* = .66, n_p_^2^ = .01. In the sensitivity analysis, where sex was added in the model, there was no evidence of a main effect of sex (*p* = .68), and still evidence of a main effect of group (*p* = .039). Removing one outlier strengthened the evidence of a main effect of Group (*p* = .015, n_p_^2^ = .10).

### Exploratory Analyses

#### Correlations between tasks and self-report measures

In line with the original EEfRT study (Treadway et al., 2009), we found evidence of a negative correlation between proportion of hard-task choices and trait anhedonia (CPAS; *r* = -.29; 95% CI *=* -0.47 to -0.10). In the Sweet Taste Test, we found evidence of positive a correlation between detection threshold and symptoms of depression (BDI-II), anhedonia (SHAPS and CPAS) and apathy (AES), see Table 2. In the JORT, to facilitate correlational analyses, individual slopes (force exerted across the four reward magnitudes) were extracted for each participant. There was no clear evidence of correlations between self-report measures and the JORT, see Table 2.

**Table 2.** Spearman’s correlations between behavioural tasks and self-report measures.

| Variable |  | JORT | EEfRT | Sweet Taste Test |
| --- | --- | --- | --- | --- |
| **SHAPS** | *R* | -.09 | -.14 | .27 |
|  | *95% CI* | -0.28 to 0.11 | -0.33 to 0.06 | 0.07 to 0.46 |
| **CPAS** | *R* | -.03 | -.29 | .22 |
|  | *95% CI* | -0.22 to 0.17 | -0.47 to -0.10 | 0.02 to 0.41 |
| **TEPS-A** | *R* | .001 | .16 | -.15 |
|  | *95% CI* | -0.20 to 0.20 | -0.04 to 0.34 | -0.34 to 0.06 |
| **TEPS-C** | *R* | -.07 | .09 | -.19 |
|  | *95% CI* | -0.26 to 0.13 | -0.11 to 0.28 | -0.38 to 0.02 |
| **BDI-II** | *R* | -.04 | -.06 | .27 |
|  | *95% CI* | -0.23 to 0.16 | -0.25 to 0.14 | 0.06 to 0.45 |
| **Apathy Scale** | *R* | .01 | -.04 | .23 |
|  | *95% CI* | -0.18 to 0.21 | -0.24 to 0.15 | 0.03 to 0.42 |

*Note.* Sweet Taste Test - mean detection threshold; EEfRT, Effort-Expenditure for Rewards Task – overall proportion of hard-task choices; JORT, Joystick-Operated Runway Task – slope of average force across four reward magnitudes; SHAPS, Snaith-Hamilton Pleasure Scale; CPAS, Chapman Physical Anhedonia Scale; TEPS, Temporal Experience of Pleasure Scale (A, Anticipatory; C, Consummatory); BDI-II, Beck Depression Inventory-II; Apathy Scale, Apathy Evaluation Scale. 95% CI, 95% Confidence Intervals; *N* = 91–101.

#### Correlations between tasks

There was evidence of a positive correlation between proportion of hard-task choices (EEfRT) and average force slopes (JORT), *r*_(101)_ = .21, 95% CI = 0.01 to 0.39, participants who were more willing to choose the hard-task option exerted more force for higher compared to lower reward magnitudes on the JORT. There was no evidence of a correlation between the EEfRT and Sweet Taste Test (*r*_(92)_ = -.02, 95% CI = -0.23 to 0.18), or the JORT and Sweet Taste Test (*r*_(92)_ = -.07, 95% CI = -0.27 to 0.13).

## Discussion

We found that the JORT was sensitive to reward, with participants exerting more force for higher reward magnitudes, but we found no evidence of a difference between the high and low anhedonia group in physical effort exerted for reward. However, compared to the low anhedonia group, the high anhedonia group displayed altered effort-based decision-making (EEfRT) and reduced reward sensitivity (Sweet Taste Test). Although the JORT did not find any effect between groups, there was an association between the EEfRT and JORT, suggesting that they may measure similar aspects of reward motivation. In this study, the EEfRT was more sensitive to changes in anhedonia scores, possibly because it involves both physical effort and cognitive decision-making.

### Reward motivation and anhedonia

In the JORT, there was no evidence of a difference between the high and low anhedonia group, both groups exerted more physical force for higher reward trials. This suggests that people with higher levels of anhedonia, at least in a non-clinical population, do not display deficits in their exertion of physical effort for reward on this task. Using a human Progressive Ratio Task, Hershenberg and colleagues (2016) reported that people with MDD had a lower breakpoint compared to healthy controls (Hershenberg et al., 2016). However, this task required cognitive effort and as the rodent Progressive Ratio Task has failed to find a similar effect in some models of depression (chronic mild stress and maternal separation) (Barr & Phillips, 1998; Shalev & Kafkafi, 2002), its translational validity has been questioned. Nevertheless, Cléry-Melin et al. (2011) found that compared to healthy volunteers, people with depression exerted less physical force to maximize monetary rewards on a handgrip apparatus (Cléry-Melin et al., 2011). Whilst these studies may seem to conflict the findings presented here, it is important to note that our study included a non-clinical population, measuring anhedonia as a varying trait, as opposed to clinical depression. Further studies are required that explore anhedonia and physical effort for reward using different contexts (e.g., real-life settings), durations (e.g., temporary vs sustained effort), and types of effort (e.g., cognitive, social, other types of physical exertion).

In the EEfRT, the high anhedonia group displayed a reduced proportion of hard-task choices, compared to the low anhedonia group. However, evidence for this difference was weak (especially after removing one outlier) and we did not find evidence of a difference between groups on 50% probability trials, as predicted in our pre-registration. Nevertheless, our exploratory analysis did replicate the original Treadway (2009) finding that higher trait anhedonia (measured using the CPAS) was associated with reduced hard-task choices on the EEfRT (Treadway et al., 2009). Interestingly, both our study and the Treadway study found an association between the EEfRT and CPAS, but failed to find a robust association between the EEfRT and SHAPS (Treadway et al., 2009; Yang et al., 2014). One possible reason for this is that the SHAPS focuses more on consummation of reward (“I would be able to enjoy my favourite meal”) whereas the CPAS also includes questions on interest/desire for future rewards (“I have had very little desire to try new kinds of foods”) (Leventhal, Chasson, Tapia, Miller, & Pettit, 2006; Rizvi, Pizzagalli, Sproule, & Kennedy, 2016), with the latter being more relevant to the EEfRT (although see Tran et al., 2020). Overall, this finding highlights the value of employing multiple anhedonia questionnaires.

There was a correlation between the JORT and EEfRT suggesting that they may measure a similar construct of motivation. However, only the EEfRT was related to anhedonia. The EEfRT examines explicit decision-making (Treadway et al., 2009) (similar to rodent choice-based tasks) where a participant must weigh up the value of two options, whereas the JORT (more similar to the Progressive Ratio Task) measures amount of physical effort exerted for reward. Speculatively, anhedonia may not be related to reduced exertion of physical effort for reward when engaged in an activity, but rather in the choice to engage in an effortful activity for reward (decision-making; Halahakoon et al., 2020). In relation to the rodent literature, these findings suggest that effort-based-choice tasks may provide a more sensitive measure of motivational impairments relevant to anhedonia than physical effort-for-reward tasks (e.g., Progressive Ratio Task). Nevertheless, it is important to consider alternative explanations including methodological differences between tasks (e.g., presence of probabilistic cues, reward sizes) and the cognitive demands of the EEfRT (Cooper et al., 2019; Whitton, Merchant, & Lewandowski, 2020).

### Reward sensitivity and anhedonia

In the Sweet Taste Test, the high anhedonia group displayed reduced sensitivity to sucrose compared to the low anhedonia group. Most previous studies using this task have compared people with clinical disorders such as MDD to healthy controls, and found conflicting results (Amsterdam et al., 1987; Berlin et al., 1998; Dichter et al., 2010). In relation to anhedonia, Bedwell and colleagues (2019) reported that sucrose sensitivity did not relate to anhedonia measures in the typical population (Bedwell, Spencer, Chirino, & O’donnell, 2019). One potential explanation for these conflicting results is how sucrose sensitivity is measured. Most studies have measured sweet taste intensity on a point scale (Bedwell et al., 2019; Dichter et al., 2010), as opposed to detection threshold (i.e., point at which sucrose can be detected) (Berlin et al., 1998). It has been suggested that point scales may not be sensitive enough to reliably detect subtle impairments (McCabe, 2018). This study suggests that anhedonia, at least in a non-clinical population, is related to reduced reward sensitivity. Future studies should examine whether this effect is specific to pleasant stimuli (e.g., sucrose) or whether it can also be observed using negative stimuli (e.g., bitter tastes), indicating a general sensitivity impairment.

Collectively, using a battery of translational tasks designed to probe different reward components (motivation, sensitivity), we support and extend preclinical findings by revealing that these components may also be dissociable in people. This is demonstrated by a lack of correlations between tasks: individuals who were less willing to choose the hard-task choice on the EEfRT did not display reduced sensitivity on the Sweet Taste Test. We also found that anhedonia was related to reduced reward motivation and reward sensitivity. Despite performance on both tasks being related to anhedonia, there was a lack of correlation between tasks. Speculatively, anhedonia may be a heterogenous symptom (i.e., there may be sub-groups of individuals who have dysfunction in different components of reward) (Thomsen, 2015; Treadway & Zald, 2013). To directly address this question, further studies (with a larger sample size) should be conducted that: (1) demonstrate that the behavioural tasks used are reliable to individual effects, and (2) employ cluster analysis on a battery of behavioural tasks (motivation, sensitivity, and learning) and self-report measures in a clinical population.

### Limitations

This study examined anhedonia in a non-clinical, predominantly student, population. There are benefits of examining non-clinical populations including the reduced likelihood of cognitive deficits, medication use and comorbidity. However, anhedonia levels in the clinical population are higher and may be qualitatively different. Therefore, future studies employing a battery of translational behavioural tasks in a clinical population are needed. Second, there was a higher proportion of males in the high anhedonia group (53%) compared to the low anhedonia group (21%), which may have influenced the differences between groups. However, including sex as a covariate in all analyses did not change the overall findings. Interestingly, few studies have investigated sex differences in anhedonia, and thus it is unknown whether the higher proportion of males in the high anhedonia group reflects a real sex difference in anhedonia (Chan et al., 2012; Xinhua Yang, Wang, Liu, Liu, & Harrison, 2020) or whether this is a result of sampling. Third, the data did not always meet the ANOVA assumptions. Whilst we attempted to address this (e.g., transforming the data), this was not always entirely successful. In these cases, analyses were conducted on the original untransformed data, which should be considered when interpreting our findings. Nevertheless, ANOVA is relatively robust to minor deviations from normality if extreme outliers are not the cause of skew (Field, 2005). Fourth, we did not include pleasantness ratings and therefore we could not assess whether there are differences in reward liking between groups. We decided not to include pleasantness ratings because this requires using point scales which may not be sensitive enough to reliably detect impairments (McCabe, 2018). For example, some studies have reported that adolescences with symptoms of depression and people with a history of depression display altered neural responses to hedonic stimuli (chocolate) compared to healthy controls, despite no evidence of a difference between groups on self-report measures of liking and intensity after tasting chocolate (McCabe, Cowen, & Harmer, 2009; Rzepa, Fisk, & McCabe, 2017). An objective measure of reward liking, which does not rely on point scales, in the human literature would be ideal to address this issue.

To summarize, this study employed a battery of behavioural tasks designed to tap into different reward components (effort-for-reward, effort-based decision-making, and reward sensitivity) in humans. Examining these different components in relation to anhedonia, we found that symptoms of anhedonia in a non-clinical population were related to changes in reward sensitivity and effort-based decision-making. Speculatively, as performance on the effort-based decision-making (EEfRT) and reward sensitivity (Sweet Taste Test) tasks did not correlate, this could suggest heterogeneity within anhedonia (Treadway & Zald, 2013). For example, some people with anhedonia may display reduced sensitivity to reward, whilst others may display altered decision-making to engage in effortful activities for reward. Further studies are required that directly address this question. If this finding is found in clinical populations, it may have important implications for assessment and treatment of anhedonia.

## Acknowledgements

The authors wish to thank Michael T. Treadway for providing us with the EEfRT.

## Ethical standards

The authors assert that all procedures contributing to this work comply with the ethical standards of the relevant national and institutional committees on human experimentation and with the Helsinki Declaration of 1975, as revised in 2008.

