## Supplementary for "Objective measures of reward sensitivity and motivation in people with high vs low anhedonia"

Supplementary Materials

#### EXPERIMENT 1: JORT as a measure of reward motivation

#### Method

##### Participants

Twenty-five participants were recruited through advertisements within the University of Bristol. Eligibility criteria were aged ≥ 18 years, fluent in English, good mental and physical health with no current or previous diagnosis of a psychiatric illness (self-reported). The study was approved by the Faculty of Biomedical Sciences Research Ethics Committee at the University of Bristol (Ref: 62301).

##### Joystick-Operated Runway Task

The JORT was originally developed as a translational measure of fear and anxiety (Perkins et al., 2009). Here, we adapted the task to measure physical effort for reward whereby participants push a force-sensing joystick to chase and catch an onscreen target for points (exchanged for money). This task is described in the main paper. The only difference between versions is that in this initial proof-of-concept study max calibrated force was calculated as peak force reached during the calibration phase. In the anhedonia study, this was changed to peak force reached during the calibration phase *or practice trials* (whichever was highest) to reduce the number of participants achieving over 75% trials and therefore accomplishing trials (120% effort trials) which were designed to be impossible. This reduced the proportion of participants achieving over 75% trials: proof-of-concept study (28% participants) and anhedonia study (6% participants).

##### Data Analysis

The following outcomes were extracted: relative average force, relative maximum force, and reaction time. For average and maximum force, mean force exerted across trials was divided by a participant’s max calibrated force to standardise performance. Trials where no force was exerted were coded as 0. Based on *a priori* criteria, participants who succeeded in over 75% trials were excluded from the analysis. This is because these participants must have achieved at least one of the trials designed to be impossible (120% effort trials) and were therefore considered to have not successfully achieved their maximum force during the calibration.

Data were exported from Python 3.6 into SPSS v24 (IBM) for analysis. Repeated measure ANOVAs (block x effort x reward) were conducted to measure relative average force, relative maximum force, and reaction time. For data assumption checks, see Experiment 2.

#### Results

Twenty-five participants aged 18 - 39 years (mean age 23.2 ± 5.3, 12 female) took part in this study. Seven participants achieved over 75% trials and were excluded from the analysis. In total, 18 participants were included in the analysis for relative average force and maximum force (mean age = 21.9, *SD* = 4.0, 7 females). For reaction time, 16 participants were included in the analysis (*N* = 2 had missing data due to no response on 0-point trials).

##### Assumptions

Residuals for some trial types were not normally distributed based on Kolmogorov-Smirnov (≤ 5/32 variables).

##### Relative Average Force

There was strong evidence of a main effect of reward magnitude, $F_{(1.10,18.76)} = 8.91$, $p = .006$, $n_p^2 = .34$. Participants exerted more force for higher reward magnitudes: 0 (*M* = 56.20, *SE* = 3.94), 10 (*M* = 63.36, *SE* = 1.58), 100 (*M* = 65.70, *SE* = 1.44) and 1000 (*M* = 69.21, *SE* = 1.61) points. Bonferroni-corrected pairwise comparisons revealed evidence of a difference between all reward magnitudes (*p*s ≤ .032), except between 0 points and 10 points ($p = .17$) and with weaker evidence of a difference between 0 points and 100 points ($p = .079$). There was weak evidence of a main effect of block, $F_{(1.0,17.0)} = 4.60$, $p = .047$, $n_p^2 = .21$. Participants exerted more force in the first block (*M* = 65.19, *SE* = 1.68) compared to the second block (*M* = 62.05, *SE* = 2.12). There was evidence of a main effect of effort, $F_{(1.62,27.59)} = 34.65$, $p < .001$, $n_p^2 = .67$. Participants exerted more force for higher effort trials: 50% (*M* = 56.11, *SE* = 1.47), 80% (*M* = 65.30, *SE* = 1.29), 100% (*M* = 67.68, *SE* = 2.21) and 120% (*M* = 65.38, *SE* = 2.47). Bonferroni-corrected pairwise comparisons revealed participants exerted less force on the 50% effort trials compared to all other effort trials (*p*s < .001) and less force on 120% effort trials compared to 100% effort trials ($p = .043$). There was no evidence of a difference between 80% and 100% effort trials (*p*s ≥ .37). There was weak evidence of a block x reward interaction, $F_{(1.85,31.5)} = 3.35$, $p = .051$, $n_p^2 = .17$. Bonferroni-corrected simple effects analysis revealed participants exerted less force in the second block compared to the first block on 0-point trials ($p = .037$), with weaker evidence of a difference between other reward magnitudes (*p*s ≥ .054). There was evidence of an effort x reward interaction, $F_{(3.06,51.95)} = 2.87$, $p = .044$, $n_p^2 = .15$, with the effect of reward being largest on 120% effort trials. There was no evidence of other interactions (*p*s ≥ .46). Re-running the analysis with 3 statistical outliers excluded weakened the evidence of a main effect of block ($p$ = .24). Re-running the analysis with all participants included ($N$ = 25) weakened the evidence of a main effect of block ($p$ = .15) and effort x reward interaction ($p$ = .75) but improved the block x reward interaction ($p$ = .007).

**Relative Maximum Force**

There was strong evidence of a main effect of reward magnitude, $F_{(1.11,\ 18.86)}$ = 9.26, $p$ = .006, $n_p^2$ = .35. Participants exerted a higher maximum force for higher reward magnitudes: 0 ($M$ = 79.30, $SE$ = 5.37), 10 ($M$ = 91.08, $SE$ = 2.53), 100 ($M$ = 94.32, $SE$ = 2.56) and 1000 ($M$ = 98.73, $SE$ = 2.94) points. Bonferroni-corrected pairwise comparisons revealed evidence of a difference in maximum force exerted between 0 and 1000 points ($p$ = .030), 10 and 100 points ($p$ = .011) and 10 and 1000 points ($p$ = .006). There was also weak evidence of a difference between 0 and 100 points ($p$ = .050) and 100 and 1000 points (p = .071) but no clear evidence of a difference between 0 and 10 points ($p$ = .12). There was evidence of a main effect of effort, $F_{(3,51)}$ = 16.38, $p$ < .001, $n_p^2$ = .49. Bonferroni-corrected pairwise comparisons revealed evidence of a difference between maximum force exerted on 50% effort trials and all other effort trials ($p$s ≤ .005) and weak evidence of a difference between 100% and 120% effort trials ($p$ = .062) but no clear evidence of a difference between other effort trials ($p$s = 1.0). There was no evidence of other main effects or interactions ($p$s ≥ .103). Re-running the analysis with one statistical outlier removed did not substantially change the findings. Re-running the analysis with all participants included ($N$ = 25) did not change the findings, but there was evidence of a block x reward interaction ($p$ = .016).

**Reaction Time**

There was strong evidence of a main effect of reward magnitude, $F_{(1.51,22.65)}$ = 8.91, $p$ = .003, $n_p^2$ = .37. Participants were quicker to respond to higher reward magnitudes: 0 ($M$ = 493 ms, $SE$ = 25), 10 ($M$ = 462 ms, $SE$ = 19), 100 ($M$ = 463 ms, $SE$ = 20) and 1000 ($M$ = 442 ms, $SE$ = 17) points. Bonferroni-corrected pairwise comparisons revealed participants were quicker to respond to 1000 points compared to other points ($p$s ≤ .022) and weak evidence of a difference between 0 and 10 points ($p$ = .050). There was no evidence of a difference between other reward magnitudes ($p$s ≥ .31). There was evidence of a main effect of block, $F_{(1.0,15.0)}$ = 16.30, $p$ = .001, $n_p^2$ = .52. Participants were quicker to respond in the first block ($M$ = 450 ms, $SE$ = 19) compared to the second block ($M$ = 481 ms, $SE$ = 21). There was evidence of a main effect of effort, $F_{(3,45)}$ = 4.76, $p$ = .006, $n_p^2$ = .24. Bonferroni-corrected pairwise comparisons revealed evidence of a difference in reaction time between 50% and 80% effort trials (mean difference = 17 ms, $p$ = .036) and weak evidence of a difference between 80% and 120% effort trials ($p$ = .058). There was no evidence of a difference between other effort trials (*p*s ≥ .17). There was evidence of an effort x reward interaction, $F_{(3.29, 49.35)}$ = 4.01, *p* =.01, $n_p^2$ = .21. There was evidence of a block x effort x reward interaction, $F_{(3.46, 51.96)}$ = 3.36, *p* = .02, $n_p^2$ = .18. Notably, including sex as a potential covariate in the model revealed evidence of a main effect of sex (*p* = .004), with males having a quicker reaction time (*M* = 431 ms, *SE* = 18; *N* = 11) compared to females (*M* = 541 ms, *SE* = 27, *N* = 5). Re-running the analysis with all participants (*N* = 22) included did not qualitatively change the findings.

#### EXPERIMENT 2: Anhedonia and reward processing

#### Method

##### Sample Size Calculation

We had sufficient resources to recruit 66 participants (33/group). An *a priori* power calculation using G*Power 3.1 (Faul, Erdfelder, Lang, & Buchner, 2007) indicated that 66 participants would provide sufficient power (.90) to detect a medium effect size of Cohen's *f* = .25 at a conventional alpha level (.05). There were two groups with four measurements. Correlations among repeated measurements was 0.50 and non-sphericity correction was 0.33. Given that we did not have any information to enable the estimation of a target effect size, the results should be interpreted with caution given the possibility that the study is underpowered.

##### Self-report measures

**Anhedonia.** The SHAPS (Snaith et al., 1995) was the primary measure of hedonic capacity used in this study. It was administered at screening (online) and re-administered during the study visit. This 14-item questionnaire asks participants to report their ability to experience pleasure in the last few days (e.g., smell of freshly baked bread, being with family). Each answer is scored on a 4-point scale ranging from 1 (strongly agree) to 4 (strongly disagree). Total scores range from 14 to 56, with higher scores indicating higher levels of anhedonia (i.e., reduced hedonic capacity). The psychometric properties of the continuous algorithm have been supported, with satisfactory test-retest reliability after 3 weeks (*r* = .70) (Franken, Rassin, & Muris, 2007), good-internal consistency (*a* = .91) (Franken et al., 2007) and good convergent validity with the Positive and Negative Affect Schedule (*r* = - .23; PANAS) (Franken et al., 2007). We originally included the SHAPS as reported in the original paper (Snaith et al., 1995), however due to poor test-retest reliability we later changed the order of responses to be consistent across questions.

The Chapman Physical Anhedonia Scale (CPAS) (Chapman, Chapman, & Raulin, 1976) measures trait anhedonia and the Temporal Experience of Pleasure Scale (TEPS) (Gard, Gard, Kring, & John, 2006) aims to dissociate consummatory and anticipatory anhedonia.

**Depression**. The Beck Depression Inventory-II (BDI-II)(Beck, Steer, & Brown, 1996) was used to measure symptoms of depression. This is a 21-item self-report questionnaire with a 4-point scale. Total scores range from 0 to 63. Higher scores indicate higher depressive symptomatology. Due to experimenter error, one response option to the 'changes in appetite' question was not available '3a. I have no appetite at all'.

**Apathy.** The Apathy Evaluation Scale (AES)(Marin, Biedrzycki, & Firinciogullari, 1991) was used to measure symptoms of apathy. This is an 18-item self-report questionnaire with a 4-point scale ranging from 1 (Not at all True) to 4 (Very True). Total scores range from 18 to 72, with higher scores indicating increased apathy.

**Effort-Expenditure for Rewards Task (EEfRT)**

The EEfRT (Treadway, Buckholtz, Schwartzman, Lambert, & Zald, 2009) is a computerised effort-based decision making task. On each trial, participants choose between an 'easy' and 'hard' option. The easy option requires 30 button presses within 7 seconds using the index finger on their dominant hand for 50p. The hard option requires 98 button presses using the little finger on their non-dominant hand within 21 seconds for 58p - £2. Participants have 5 seconds to make a choice or they are randomly assigned to the easy or hard option for that trial. Each trial also contains a probability cue (12%, 50%, 88%; visible to the participant) which indicates the probability of winning money if they successfully complete that trial (i.e., "win trials"). The probability cue applies to both the easy and hard task choice, with an equal proportion across the experiment. Participants are informed that two of these "win trials" may be paid to them at the end of the experiment (performance-based pay). The task takes 20 minutes to complete the same pre-randomised order of trials.

Prior to the task, participants complete three trials to assess their motor ability (i.e., finger tapping rate) where they press a button as fast as possible with the little finger on their non-dominant hand for 21 seconds. This is followed by four practice trials.

The only difference between the original task and this version is that we modified the payment: the easy trial was always 50p and the hard trial was up to £2 (original task: $1.00 easy trial, $1.24 to $4.30 hard trial). Nevertheless, the proportion of money on offer on each trial is approximately the same as the original task.

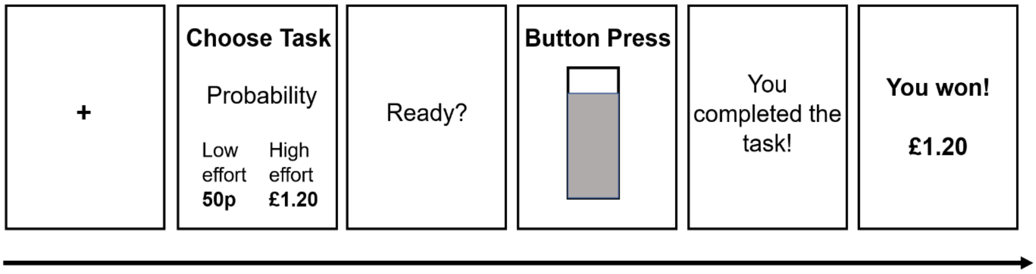


**S1.** Example trial on the Effort-Expenditure for Rewards Task (EEfRT): fixation cross (1 s), choice period where the participant is informed about the probability of the trial being a 'win trial' and reward magnitude of the hard task (5 s), 'Ready' screen (1 s), button pressing (7 s easy task or 21 s hard task), feedback as to whether the participant successfully completed the task followed by feedback (Treadway et al., 2009).

#### Data Analysis

Data were checked for normality (Kolmogorov-Smirnov), homogeneity of variances (Levene's test) and potential outliers (scores 3 times greater than the interquartile range). If either ANOVA assumption was violated and transformed data met the assumptions, data analysis was done on transformed data, otherwise analysis was conducted on non-transformed data. Greenhouse-Geiser corrections were applied where Mauchly's Test of Sphericity was $p < .05$. Data is presented with all participants retained in the analysis. Where removal of outliers qualitatively changed the findings, results will also be reported with outliers excluded. *Post-hoc* comparisons were Bonferroni-corrected for multiple comparisons.

### Results

**S2.** Distribution of scores on self-report measures

| Variable | Number | Mean | *SD* |
|---|---|---|---|
| **SHAPS** | 101 | 21.4 | 5.6 |
| **Chapman Physical Anhedonia Scale** | 100 | 12.6 | 7.3 |
| **TEPS - Anticipatory Subscale** | 100 | 45.6 | 7.2 |
| **TEPS - Consummatory Subscale** | 101 | 36.6 | 7.0 |
| **Beck Depression Inventory (BDI-II)** | 100 | 8.9 | 6.8 |
| **Apathy Evaluation Scale (AES)** | 100 | 28.6 | 6.1 |

*Note.* SHAPS, Snaith-Hamilton Pleasure Scale; TEPS, Temporal Experience of Pleasure Scale; SD, standard deviation. Missing data ($N = 1$) for CPAS, TEPS, BDI-II and AES where a participant either recorded more than one response to a question or did not answer all questions.

**S3.** Correlations between self-report measures

| | SHAPS | CPAS | TEPS - ANT | TEPS - CON | BDI-II | AES |
|---|---|---|---|---|---|---|
| **SHAPS** | | .43** | -.58** | -.58** | .29** | .49** |
| **CPAS** | | | -.52** | -.67** | .28** | .48** |
| **TEPS - ANT** | | | | .65** | -.26** | -.42** |
| **TEPS - CON** | | | | | -.27** | -.53** |
| **BDI-II** | | | | | | .47** |

*Note.* Spearman's rho, *N* = 99 - 101, * $p < .05$, ** $p < .01$. Anhedonia scales: SHAPS, CPAS, TEPS - ANT, TEPS – CON. Depression scale: BDI-II. Apathy scale: AES.

#### Effort-Expenditure for Rewards Task

In the first 50 trials, the mean percent of trials completed was 97.72% (*SD* = 4.79, range = 72 – 100%). Additionally, we checked the number of participants who only chose the easy or hard task to confirm that these participants did not drive the findings. In total, one participant only chose the easy task and there were no participants who only chose the hard task. Removal of this participant weakened the evidence of a main effect of group (*p* = .051).

**Assumptions**. Some of the residuals were not normally distributed and homogeneity of variances was not met for low probability trials.

**Reaction time.** There was strong evidence of a main effect of probability, $F_{(2,126)} = 17.91$, $p < .001$, $n_p^2 = .22$, see S4. Participants were quicker to make a choice when there was a lower probability of winning money: 12% (*M* = 1.72 seconds, *SE* = .06), 50% (*M* = 1.93 seconds, *SE* = .07) and 88% probability trials (*M* = 1.92 seconds, *SE* = 0.07). Bonferroni-corrected pairwise comparisons revealed evidence of a difference between all probability levels (*p*s < .001), except between medium and high probability trials (*p* = 1.0). There was no evidence of other main effects or interactions (*p*s ≥ .21).

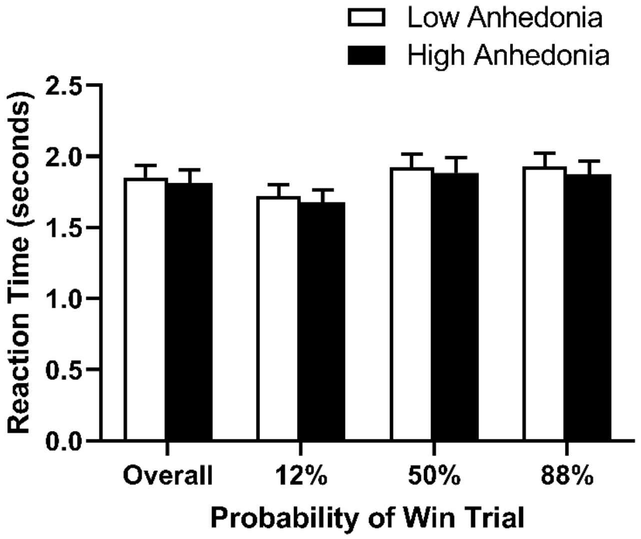


**S4**. Mean reaction time across different levels of probability for the high ($N$ = 32) and low ($N$ = 34) anhedonia group. Error bars represent *SEM*.

**Finger tapping speed.** There was no evidence of a difference between high ($M$ = 98.54, $SD$ = 9.49) and low ($M$ = 97.23, $SD$ = 9.92) anhedonia groups in finger tapping speed prior to the task $t_{(64)}$ = -.55, $p$ = .58, which may suggest any differences between groups cannot be attributed to differences in motor ability.

**Joystick-Operated Runway Task**

**Assumptions.** Some of the residuals were not normally distributed (2/8 variables).

**Maximum force.** There was strong evidence of a main effect of reward $F_{(1.47,88.08)}$ = 36.85, $p$ < .001, $n_p^2$ = .381, see S5. Participants exerted a higher maximum force for higher reward magnitudes: 0 points ($M$ = 73.90, $SE$ = 3.00), 10 points ($M$ = 85.86, $SE$ = 2.38), 100 points ($M$ = 91.16, $SE$ = 1.91) and 1000 points ($M$ = 94.08, $SE$ = 1.89). Bonferroni-corrected pairwise comparisons revealed strong evidence of a difference between all reward magnitudes (*p*s ≤ .006). There was no evidence of a main effect of group ($F_{(1, 60)}$ = .49, $p$ = .49, $n_p^2$ = .008; see S5) or reward x group interaction ($F_{(1.47,88.08)}$ = .67, $p$ = .47, $n_p^2$ = .01). In the sensitivity analyses, where sex was added to the model, there was still no evidence of a main effect of group ($p$ = .81), sex ($p$ = .24), or reward x group interaction ($p$ = .50).

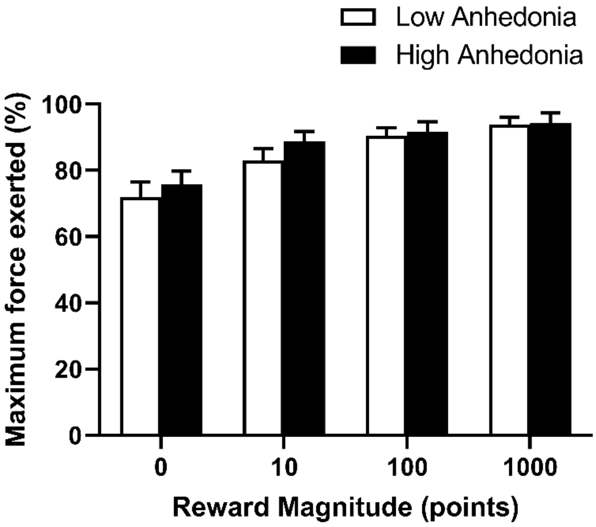


**S5.** Maximum force exerted across different reward magnitudes on the JORT for the high (*N* = 29) and low (*N* = 33) anhedonia group. Error bars represent *SEM*.

**Reaction time.** There was strong evidence of a main effect of reward, $F_{(1.18,70.73)}$ = 13.90, $p$ < .001, $n_p^2$ = .19, see S6. Participants were quicker to respond to higher reward magnitudes: 0 points (*M* = 520 ms, *SE* = 15), 10 points (*M* = 477 ms, *SE* = 11), 100 points (*M* = 479 ms, *SE* = 11) and 1000 points (*M* = 468 ms, *SE* = 11). Bonferroni-corrected pairwise comparisons revealed evidence of a difference between all reward magnitudes (*p*s ≤ .024), except between 10 and 100 points (*p* = 1.0). There was no evidence of a main effect of group ($F_{(1,60)}$ = .86, *p* = .36, $n_p^2$ =.01; see S6) or reward x group interaction ($F_{(1.18,70.73)}$ = .049, *p* = .86, $n^2_p$ = .001). There was one outlier, which when removed did not change the findings but did improve the reward x group interaction, $F_{(1.97,116.10)}$ = 2.30, *p* =.106, $n_p^2$ = .04. In the sensitivity analysis, where sex was added to the model, there was evidence of a main effect of sex, $F_{(1,59)}$ = 13.98, *p* < .001, $n_p^2$ = .19, with males being quicker to respond (*M* = 433 ms, *SE* = 17; *N* = 21) than females (*M* = 515 ms, *SE* = 12; *N* = 41). There was still no evidence of a main effect of group (*p* = .72) or reward x group interaction (*p* = .79).

Re-running all analyses (average force, maximum force, and reaction time) as full 2*4*4 ANOVAs still did not reveal any evidence of a main effect of group or interactions with group (*p*s ≥ .18).

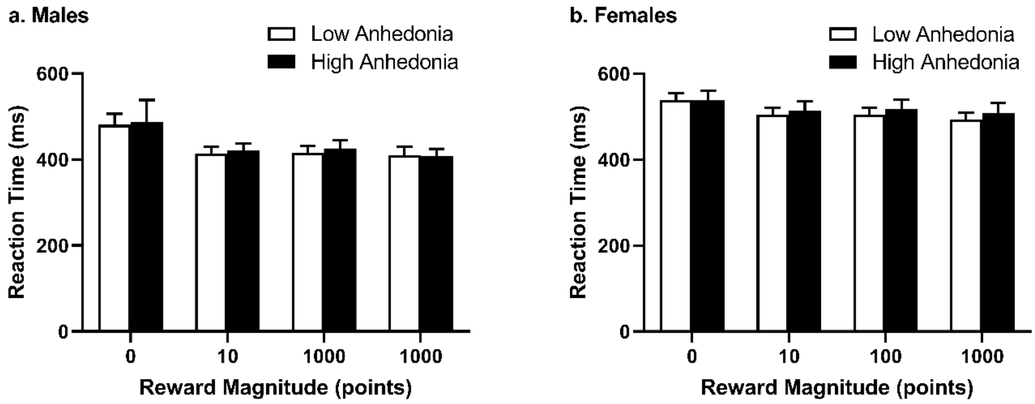


**S6.** Reaction time (ms) across different reward magnitudes on the JORT for the high and low anhedonia group. As there was a sex difference in reaction time on the JORT ($p < .001$), data are presented separately for (a) males ($N = 6$ low, 15 high anhedonia) and (b) females ($N = 27$ low, 14 high anhedonia). Error bars represent *SEM*.

##### Sweet Taste Test

This task was originally run in an air-conditioned room. Malfunction of the air-conditioning system part way through the study meant that some participants completed the study with air-conditioning, whilst others did not. As temperature may effect taste sensitivity, we ran an exploratory analysis including this as a control variable. Including this control variable in a univariate ANOVA, still revealed evidence of a main effect of group ($p = .015$).

**S7.** Spearman correlations between EEfRT (hard-task choices at each probability level) and questionnaires

| Variable | HC 88% probability | HC 50% probability | HC 12% probability | Number |
|---|---|---|---|---|
| **SHAPS** | -.06 | -.12 | **-.21*** | 101 |
| **CPAS** | **-.22*** | **-.25*** | **-.23*** | 100 |
| **TEPS – A** | .11 | .15 | **.21*** | 100 |
| **TEPS – C** | .01 | .09 | .10 | 101 |
| **BDI-II** | -.02 | -.03 | -.07 | 100 |
| **Apathy** | .001 | -.01 | -.13 | 100 |

*Note.* HC, hard-task choices; SHAPS, Snaith-Hamilton Pleasure Scale; CPAS; Chapman Physical Anhedonia Scale; TEPS, Temporal Experience of Pleasure Scale (A, anticipatory; C, consummatory), BDI-II, Beck Depression Inventory II; Apathy, Apathy Evaluation Scale. Spearman's rho, *** *p* < .05.**

#### Anhedonia and task specificity

For the Sweet Taste Test, re-running the analysis with performance on the EEfRT (proportion of hard-task choices) included as a covariate weakened the main effect of group on detection threshold ($p$ = .047). For the EEfRT, rerunning the analysis with performance on the sweet taste test (detection threshold) as a covariate weakened the evidence for a main effect of group on proportion of hard-task choices ($p$ = .070). This supports the exploratory analysis which suggests a stronger relationship between the SHAPs (used to group individuals into high vs low anhedonia) and the sweet taste test, whereas the EEfRT had a stronger relationship with the CPAS.
